# The association of objectively and subjectively measured *modifiable* lifestyle factors with internalizing problems: The role of genetic confounding and shared method variance *bias*

**DOI:** 10.1101/2024.11.02.24316385

**Authors:** Yingzhe Zhang, Karmel Choi, Leonard Frach, Elise Robinson, Tian Ge, Jean-Baptiste Pingault, Henning Tiemeier

## Abstract

**Background:** Sleep duration and physical activity have been associated with internalizing problems, such as depression, in observational studies. However, genetic confounding and measurement error may introduce bias. We assessed genetic confounding in the associations of sleep duration and physical activity with internalizing problems using both device-based and questionnaire assessments to estimate shared genetic risk across different assessment methods in adolescents.

**Methods:** In this preregistered study embedded in the Adolescents Brain Cognitive Development cohort, we included unrelated European adolescents with both self-reported and Fitbit-measured sleep duration devices data (N = 2283) and data on moderate to vigorous physical activity (days/week, N = 2772). Adolescents had a mean age of 12-year-old (SD = 0.65), with roughly 48% female. The internalizing problem scores were derived from self-reports of the Brief Problem Monitor A genetic sensitivity analysis was conducted to assess genetic confounding by combining polygenic scores and molecular-based heritability estimates of internalizing problems.

**Results:** Longer sleep duration was associated with lower internalizing problems using both self-reported (-0.15-SD, 95% CI, -0.19 to -0.11-SD) and objective (-0.10-SD, 95% CI, -0.15 to -0.06-SD) assessments. More frequent moderate/vigorous physical activity was associated with lower internalizing problems using both self-reported (-0.09-SD 95% CI, -0.13 to -0.05-SD) and device-based (-0.06-SD, 95% CI, -0.09 to -0.01-SD) assessments. A higher internalizing polygenic score was associated with more internalizing problems and shorter self-reported sleep duration but not device-based assessed sleep duration. Substantial genetic confounding (81%) was found between self-reported sleep duration and internalizing problems, predominantly among boys. No clear evidence of genetic confounding was found in the association between device-based measured sleep duration and internalizing problems. Similarly, no genetic confounding was observed for measures of physical activity in relation to internalizing problems.

**Conclusion:** The observed negative relationship between reported child sleep duration and internalizing problems may be partly due to genetic confounding, particularly among boys. This genetic influence likely captured some measurement error (i.e., shared method variance) in associations where exposure and outcome were assessed by self-reports. Observational studies relying on self-reports may overestimate the impact of adolescent sleep duration on internalizing problems, especially among boys. Questionnaire assessment of physical activity was less impacted.

## Introduction

Sufficient sleep duration and adequate physical activity are commonly considered modifiable lifestyle behaviors that can alleviate adolescent mental health, especially internalizing problems,^1,2^ encompassing anxiety, depression, and somatic pain. Observational studies have linked shorter sleep duration^3-5^ and less physical activity^6,7^ with depression in adolescents, but meta-analyses on intervention studies have showed inconsistent results.^8,9^ These discrepancies may be partly explained by the underlying shared genetic architectures for lifestyle risk factors, such as sleep and physical activity, and internalizing problems, which are both known to be influenced by genetic factors.^10,11^ Genome-wide association studies (GWAS) of physical activity and sleep duration have identified overlapping genetic variants associated with depression.^12,13^ Thus, there could be shared genetic risk (i.e., genetic confounding) as a common cause that creates associations between adolescent lifestyle behaviors and internalizing problems in the absence of a causal effect. Genetic factors affecting sleep or physical activity might also influence internalizing problems through biological pathways such as dopamine binding and synaptic neurotransmission.^14,15^ Shared genetic risk factors are prevalent in behavioral research,^16,17^ complicating causal interpretations and potentially biasing assessments of environmental influences.^18,19^ Despite this, the role of genetic confounding in the relationship between lifestyle factors and internalizing problems remains underexplored.

Observational studies on sleep, physical activity and mental health may suffer from measurement error (i.e., information bias) due to reliance on self-reported questionnaire data for both exposure and outcome measures.^20,21^ Such shared method variance bias can typically inflate observed phenotypic associations.^22^ The shared genetic risk may be compounded by information bias because a genetic predisposition for depression may influence how lifestyle factors and internalizing problems are reported. Subjective measures are susceptible to errors stemming from memory, perception, and social desirability,^23^ which may all be partially genetically influenced. Longer sleep duration and more physical activity are typical from self-reports compared to objective records.^24-27^ Specifically, internalizing problems could be associated with recall bias and might directly influence perception of lifestyle factors.^28^ Importantly, depression is associated with greater discrepancy in subjective and objective sleep duration.^29,30^Adolescents with a higher genetic risk of internalizing problems may be more likely to report that sleep or physical activity level is poor. To mitigate these biases, objective measures such as wearable devices (e.g., accelerometers, Fitbit) bypass self-report biases and may therefore enhance accuracy in assessing sleep and physical activity.^31^

Given the public health importance of supporting adolescent mental health through behavioral interventions, it is crucial to i) dissect observed associations between lifestyle factors and mental health outcomes into potential genetic confounding and residual associations and ii) test whether observed associations depend on assessment methods. Our study aims to quantify the extent of genetic confounding in the observed associations of adolescent sleep duration and physical activity with internalizing problems, separately for questionnaire and device-based assessments of lifestyle factors. We hypothesized that self-reported lifestyles would be more influenced by genetic confounding in their associations with internalizing problems compared to device-based measures.

## Methods

To make our decisions transparent, we pre-registered our study hypotheses, detailed methods and procedures, and the complete data analysis plan using the Open Science Framework repository (https://osf.io/jck54) prior to data analysis.

### Study population

We used data from the Adolescent Brain Cognitive Development (ABCD Release 5.0) Study, conducted across the United States. ABCD has followed 11875 adolescents since the ages of 9-10.^32,33^ Because polygenic scores (PGS) were derived from GWAS of sleep, physical activity, internalizing problems and depression in European ancestry populations, we only included genetically unrelated adolescents of genetically identified European ancestry. In the separate analyses for the sleep duration and physical activity, we included two analytical samples with complete data in genotyping, device-based measured exposures, and self-reported exposures (N = 2253 adolescents for sleep duration and N = 2772 for physical activity).

### Sleep duration and physical activity assessment

All exposures were selected from the two-year follow-up when the participants were around 12-year-old due to availability of both self-report and device-based measures at this wave. For sleep duration, adolescents reported sleep time in hours with the Youth Munich Chronotype Questionnaire. The included adolescents were also monitored with a Fitbit device for sleep duration. For comprehensive assessment of sleep, we included adolescents with both weekday and weekend Fitbit measured records. For both assessments, we filtered out the records of sleep duration of less than 5 hours and more than 13 hours to exclude outliers that may likely reflect measurement error.

For physical activity measures, in the same assessment wave, adolescents were asked specifically “During the past 7 days, on how many days were you physically active for a total of at least 60 minutes per day?”. The included adolescents were also monitored with a Fitbit device for moderate/vigorous physical activity time (in minutes per day). We transformed this device-based measured physical activity to the number of days of more than 60 minutes moderate/vigorous physical activity to be consistent with the self-reported measure. Again, we included adolescents with both weekday and weekend assessments. We also excluded poor quality records with wearing time less than 600 minutes of non-sleep time or no heart rate more than 30 minutes per day. The mean of device wearing days was 14.9 days (SD = 9.3) for sleep duration and 12.5 days (SD = 8.4) for physical activity.

### Outcome assessment

The Brief Problem Monitor (BPM)^34^ instrument completed by adolescents at the three-year follow-up wave (i.e., one-year after lifestyles assessments) was used as the outcome. This questionnaire is designed to quickly obtain reliable responses. Internalizing problems were defined as the mean scores of 6 items (such as anxious and depressive feelings, see Supplementary Methods). As a secondary outcome measure, we used internalizing problems reported by parents with the Achenbach Child Behavior Checklist (CBCL) 6/18 at three-year follow-up wave; this allowed us to examine associations between exposure and outcome assessed by different reporters. The internalizing scale includes 32 items from anxious/depressed, somatic complaints, and withdrawn/depressed subscales. Each item in both instruments is rated from 0 to 2. Higher scores indicate more internalizing problems in both questionnaires.

### Covariate assessment

Age at exposure assessments, sex, family income, and parental education measured at baseline were included as confounders. Study sites was used as a clustered variable with random effect in the associations of child physical activity or sleep time with internalizing problems. The top 10 principal components (PCs) were additionally included as covariates to account for residual confounding by genetic ancestry in associations of PGS with phenotypes. The PGSs were pre-residualized for top 10 PCs in genetic confounding analyses.

### Missingness

For outcome variables, we imputed the missing outcome values using multiple imputation by chained equations (MICE), only if prior BPM or CBCL measures in the baseline or one-year follow-up were available, to improve precision in analyses.^35^ We also imputed the missing covariates, including age at exposure assessments, sex, family income, and parental education measured at baseline, by MICE. We used predictive mean matching method for imputations of both covariates and outcome variables.

### Genetic information

Detailed information about genotyping in ABCD has been described.^36,37^ Briefly, we extracted 7,989,528 genetic variants with minor allele frequency >0.01, SNP-level call rate >0.98, and Hardy–Weinberg equilibrium p >1e-10 based on the imputed genotyping data. We identified adolescents of European ancestry using the first 6 PCs with random forest models and unrelated individuals based on PC-AiR.^37^ After removing 36 adolescents with PC outliers within European ancestry population, we retained 4470 unrelated adolescents of European ancestry. First, we calculated a polygenic score of internalizing factors as an estimate of the genetic likelihood to develop internalizing problems. Genome-wide internalizing PGS was derived based on the summary statistics of the internalizing factor from Genomic Structural Equation Modelling from a previous study.^38^ This internalizing factor leveraged the GWAS summary statistics from depression, anxiety and post-traumatic stress disorder. We used internalizing PGS in the main analyses to match with the outcome measure of internalizing problems. Additionally, for sensitivity analyses, we generated the depression PGS based on summary statistics from a depression GWAS (n=500,199 individuals).^39^ Although these GWAS summary statistics are based on adults, previous research suggested a robust overlap between salient genetic factors in adolescents and adults.^40^ The SNP weights for PGS construction were computed using PRS-CS software, a Bayesian scoring method which places a continuous shrinkage (CS) prior on SNP effect sizes.^41^ European samples from the 1000 Genomes Project Phase 3v5 reference panel were used to model linkage disequilibrium among variants. We then used PLINK to calculate polygenic scores for individuals in the target ABCD sample by summing all included variants weighted by the inferred posterior effect size for the effect allele. Second, the heritability of internalizing problems was calculated in the unrelated European sample (N = 4062) using the genomic-relatedness-based restricted maximum-likelihood approach implemented in GCTA.^42^ We log-transformed the mean internalizing scores to achieve normality in the GCTA analyses.

### Statistical analysis

Demographic variable distributions of the included analytical samples and overall unrelated European adolescents in ABCD were analyzed to demonstrate potential sample selection. In our analyses, PGSs of both internalizing factor and depression, lifestyle factors (i.e., sleep duration and physical activity) and child internalizing problems were standardized to mean 0 and standard deviation (SD) 1 prior to residualizing to facilitate interpretation and comparisons.

For main analyses, first, we examined the phenotypic associations between lifestyle factors and internalizing problems. To this end, we tested the association between both the self-reported and device-based measured sleep duration and physical activity with self-reported internalizing problems using mixed-effect linear regressions. These analyses were adjusted for age at lifestyle assessment, sex at birth, parental education, and family income and included study site as a random effect. Additional analyses using parent reported internalizing problems were conducted. Second, we quantified the associations of internalizing PGS with the lifestyle factors and internalizing problems using mixed linear regressions adjusting for age, sex and top 10 PCs and clustering on study sites. We conducted association analyses using the PGS of internalizing factor among 2084 individuals with complete data jointly in both sleep duration and physical activity. Sex-stratified analyses were performed to identify potential sex differences.

Third, we employed the Genetic Sensitivity (Gsens) framework, based on structural equation modeling,^43^ to assess genetic confounding in the relationships between sleep duration or physical activity and internalizing problems. In the Gsens framework, the internalizing PGS is combined with internalizing SNP-based heritability to produce an estimate of genetic confounding that accounts for the fact that a PGS typically only capture a fraction of the corresponding SNP-heritability. We residualized the outcome measures for age, sex, parental education, family income, and included study site as a random effect. The GsensY framework, which relies on the polygenic score and the heritability of the outcome measure (here internalizing), was selected as it is less sensitive to amplification bias potentially resulting from unobserved nongenetic confounders.^43^ In each Gsens model, we residualized the PGS on age, sex and top 10 PCs to account for potential population stratification and clustered analyses by study sites. This approach was applied separately to both objective and subjective assessments of lifestyle factors for comparison.

Sex stratified follow-up analyses were performed given the sex differences in PGS-phenotypes associations. Furthermore, a moderate non-linear association between reported sleep duration and internalizing problems was observed. Thus, we conducted a sensitivity analysis with Gsens restricting sleep duration to values between 7 to 11 hours within a linear distributed range to meet the linearity assumption of the structural equation model. Further sensitivity analyses were conducted using the depression PGS to determine if the genetic risks associated with depression reflect consistent patterns with broader internalizing genetic risks.

## Results

### Study population

We included 2,253 adolescents in the sleep duration analyses and 2,772 adolescents in the physical activity analyses. As showed in supplementary Table 1, included samples were 12-year-old on average (SD = 0.65) when the lifestyles were assessed at two-year follow-up wave and roughly 48% were female. Among these adolescents, 48% had parents with education above graduate level and 56% came from families with an annual income above $100,000. The mean sleep duration was 9.3 hours (SD = 1.0) measured by self-report and 7.6 hours (SD = 0.5) measured by Fitbit device, with a correlation of 0.38. The mean days with more than 60 minutes of moderate or vigorous physical activity per week was 4.1 days (SD = 2.1) measured by self-report and 1.5 days (SD = 1.7) measured by device, with a correlation of 0.22. The mean internalizing item score reported by adolescents was 0.34 (SD = 0.4). We compared the distributions of demographic variables in the analytical sample of participants with genotyping data and both self-reported and device-based exposure measures to the total unrelated European sample in ABCD. Overall, characteristics were similar although our analytical sample had a higher socio-economic status. Additionally, the socio-demographic and device wearing time between boys and girls in our analytical sample were similar. As expected, girls reported higher internalizing problem scores. While self-reported sleep duration was similar between genders, the device-based measure indicated that girls had longer sleep duration compared to boys (Supple. Table2).

### Phenotypic associations

Adjusting for socio-demographic covariates, longer sleep duration and more days of moderate to vigorous physical activity were both associated with lower internalizing problems using self-reported and device-based measured lifestyle factors (Figure 1). Specifically, each additional SD of child-reported sleep duration was associated with a 0.15-SD (95% CI: 0.11 to 0.19-SD) lower internalizing problem score; each additional SD of device measured sleep duration was associated with a 0.10-SD (95% CI, 0.06 to 0.15-SD) lower internalizing problem score. Similarly, each additional SD of child-reported moderate to vigorous physical activity (days/week) was associated with a 0.09-SD (95% CI, 0.05 to 0.13-SD) lower internalizing problem score and each additional SD of device-assessed physical activity (days/week) was associated with a 0.06-SD (95% CI, 0.01 to 0.09-SD) lower internalizing problem score. For both lifestyle measures, the self-reported measure was more strongly associated with internalizing problems than the device-based assessments. In sex-stratified analyses, the estimated coefficients were larger among girls than among boys for both self-reported lifestyle factors and device assessed sleep duration (Supple. Figure 1). When using secondary outcome reported by parents, the association of sleep duration and physical activity with internalizing problems became non-significant for sleep duration (Supple. Figure 2).

**Figure 1.**
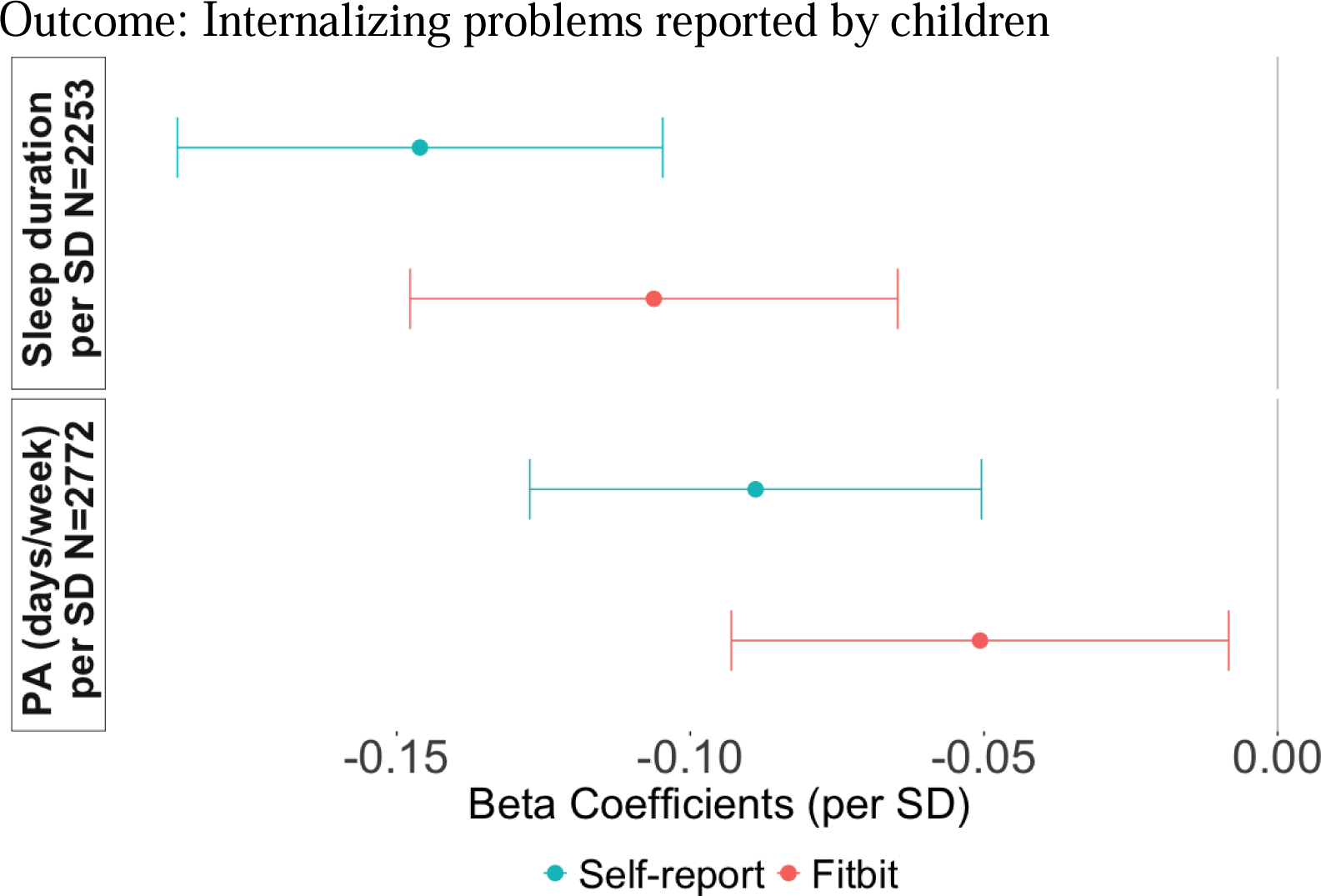
phenotypic associations between modifiable risk factors and internalizing problems reported by children. PA: Physical activity. All models were adjusted for sex, age, family income and parental education, clustered with study sites. Note: The correlation between self-reported PA and Fitbit measured PA is 0.22. The correlation between self-reported sleep duration and Fitbit measured sleep duration is 0.38.

**Figure 2.**
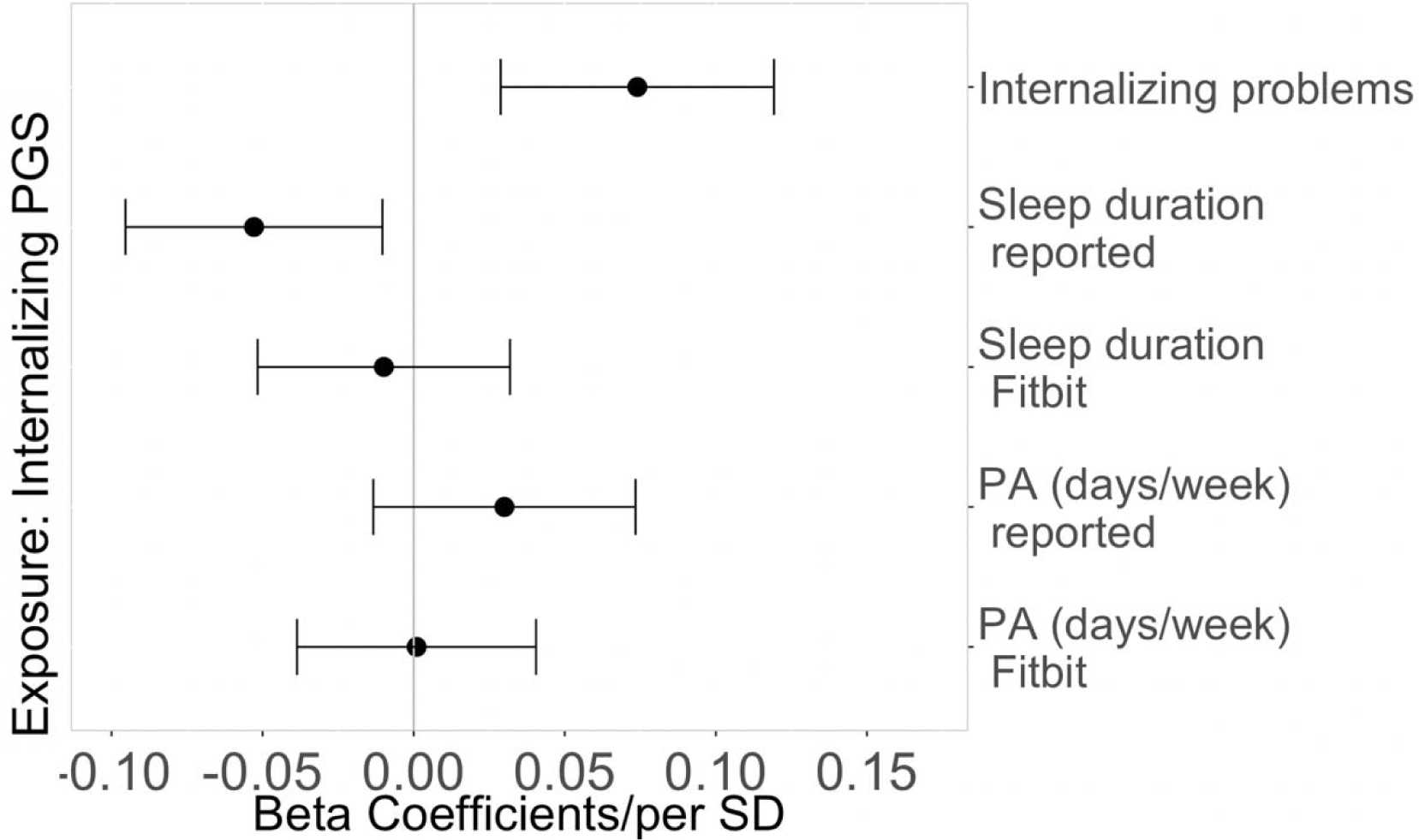
PGS of internalizing problems associations between modifiable risk factors and internalizing problems. The overlapping participants of sleep and physical activity samples (N = 2084) was included. All models were adjusted for age, sex, top 10 PCs and clustered with study sites.

### Genetics-phenotype associations

In genetic association analyses (Figure 2), we observed that a higher internalizing PGS was associated with increased internalizing problems (beta = 0.07, 95% CI 0.03, 0.12) and shorter self-reported sleep duration (beta = -0.05, 95% CI -

0.10, -0.01), but not with device-based sleep duration (beta = -0.01, 95% CI -0.05, 0.03). There was no association between internalizing PGS and either self-reported (beta = 0.03, 95% CI - 0.01, 0.07) or device measured physical activity (beta = 0.001, 95% CI -0.386, 0.041). The association of internalizing PGS with lifestyle factors and internalizing problems may be moderated by sex at birth, although the differences were not statistically significant (Supple. Figure 4). In sensitivity analyses using depression PGS, which moderately correlated with internalizing PGS (r = 0.46), we found similar patterns (Supple. Figure 3; Supple. Table 3). However, depression PGS was also linked to reduced device assessed physical activity (beta = - 0.06, 95% CI -0.10, -0.02, see Supple. Figure 3 and Supple. Table 3). Also, a pronounced sex difference (p=0.01 for interaction term of PGS and sex) was observed between the association between depression PGS and child-reported sleep duration: a significant negative association was found among boys, while no such association was detected among girls (Supple. Figure 5).

### Genetic confounding

In the 4062 unrelated European sample in the ABCD cohort, the molecular-based heritability of the internalizing problem score was 15.9% (SE = 0.08). The heritability of the internalizing problems among the 2161 boys was 11.7% (SE=0.16) and among the 1901 girls it was 25.0% (SE=0.18). These heritability estimates were incorporated into the Gsens framework with PGS.

In the analyses using Gsens models to assess genetic confounding using internalizing PGS, 81% of the association between self-reported child sleep duration and internalizing problems was attributed to genetic confounding. There were marked differences by sex: sex-stratified analyses showed that genetic confounding fully explained this association of self-reported sleep with internalizing problems in boys, and no clear evidence of genetic confounding of this association in girls (Table 1). In contrast, the relationship between device assessed sleep duration and internalizing problems showed less clear evidence of genetic confounding, regardless of sex.

**Table 1.** Genetic confounding between child sleep duration and internalizing problems.

| **Reported Sleep – INT (N = 2253)** |  |  |  | **Fitbit Sleep – INT (N = 2253)** |  |  |  |
| --- | --- | --- | --- | --- | --- | --- | --- |
|  | beta | 95% CI | p |  | beta | 95% CI | p |
| Total association | -0.147 | -0.188, -0.106 | <0.001 | Total association | -0.103 | -0.145, -0.062 | <0.001 |
| Residual association | -0.028 | -0.146, 0.089 | 0.64 | Residual association | -0.069 | -0.163, 0.025 | 0.15 |
| Genetic confounding | -0.119 | -0.228, -0.010 | 0.03 | Genetic confounding | -0.035 | -0.122, 0.052 | 0.43 |
| **Reported Sleep – INT in Boys (N = 1176)** |  |  |  | **Fitbit Sleep – INT in Boys (N = 1176)** |  |  |  |
|  | beta | 95% CI | p |  | beta | 95% CI | p |
| Total association | -0.128 | -0.179, -0.076 | <0.001 | Total association | -0.065 | -0.122, -0.008 | 0.03 |
| Residual association | 0 | NA | NA | Residual association | -0.074 | -0.212, 0.065 | 0.30 |
| Genetic confounding | -0.128 | -0.179, -0.076 | <0.001 | Genetic confounding | 0.008 | -0.121, 0.137 | 0.90 |
| **Reported Sleep – INT in Girls (N = 1077)** |  |  |  | **Fitbit Sleep – INT in Girls (N = 1077)** |  |  |  |
|  | beta | 95% CI | p |  | beta | 95% CI | p |
| Total association | -0.181 | -0.240, -0.122 | <0.001 | Total association | -0.138 | -0.197, -0.079 | <0.001 |
| Residual association | -0.028 | -0.225, 0.168 | <0.001 | Residual association | -0.037 | -0.214, 0.140 | 0.68 |
| Genetic confounding | -0.153 | -0.341, 0.036 | 0.11 | Genetic confounding | -0.101 | -0.271, 0.069 | 0.24 |

Note: Internalizing PGS and internalizing problems heritability were used in models. Internalizing PGS was residualized on age, sex, top 10 PCs and clustered with study sites, and exposures were residualized on age, sex, family income, parental education and clustered with study sites. Both sleep duration (hours) and internalizing problems score were scaled to mean 0 and standard deviation 1.

For physical activity, no genetic confounding was detected in the associations with internalizing problems, as evidenced by overlapping 95% confidence intervals and non-significant p-values (Table 2). No genetic confounding was found in both boys and girls. The association between self-reported physical activity and internalizing problems largely remained after accounting for genetic confounding. The association between device assessed physical activity and internalizing problems was only marginally significant, making accurate estimation of genetic confounding challenging.

**Table 2.** Genetic confounding between child physical activity and internalizing problems.

| **Reported PA – INT (N = 2772)** |  |  |  | **Fitbit PA – INT (N = 2772)** |  |  |  |
| --- | --- | --- | --- | --- | --- | --- | --- |
|  | beta | 95% CI | p |  | beta | 95% CI | P |
| Total association | -0.100 | -0.137, -0.063 | <0.001 | Total association | -0.049 | -0.087, -0.012 | 0.01 |
| Residual association | -0.072 | -0.140, -0.004 | 0.04 | Residual association | 0.012 | -0.063, 0.087 | 0.75 |
| Genetic confounding | -0.028 | -0.088, 0.032 | 0.36 | Genetic confounding | -0.058 | -0.124, 0.01 | 0.07 |
| **Reported PA – INT in Boys (N = 1176)** |  |  |  | **Fitbit PA – INT in Boys (N = 1176)** |  |  |  |
|  | beta | 95% CI | p |  | beta | 95% CI | p |
| Total association | -0.074 | -0.132, -0.016 | 0.01 | Total association | -0.074 | -0.131, -0.017 | 0.01 |
| Residual association | -0.166 | -0.388, 0.057 | 0.14 | Residual association | -0.075 | -0.215, 0.065 | 0.30 |
| Genetic confounding | 0.091 | -0.124, 0.307 | 0.41 | Genetic confounding | 0.001 | -0.130, 0.132 | 0.99 |
| **Reported PA – INT in Girls (N = 1077)** |  |  |  | **Fitbit PA – INT in Girls (N = 1077)** |  |  |  |
|  | beta | 95% CI | p |  | beta | 95% CI | p |
| Total association | -0.125 | -0.184, -0.066 | <0.001 | Total association | -0.034 | -0.094, 0.025 | 0.26 |
| Residual association | -0.117 | -0.291, 0.058 | 0.19 | Residual association | / | / | / |
| Genetic confounding | -0.009 | -0.178, 0.161 | 0.92 | Genetic confounding | / | / | / |

Note: Internalizing PGS and internalizing problems heritability were used in models. Internalizing PGS was residualized on age, sex, top 10 PCs and clustered with study sites, and exposures were residualized on age, sex, family income, parental education and clustered with study sites. Both physical activity (days/week) and internalizing problems score were scaled to mean 0 and standard deviation 1.

In sensitivity analyses using depression PGS within the Gsens framework, similar patterns were observed. However, genetic confounding was detected in the association between sleep duration and internalizing problems among boys for both self-reported and device assessed sleep duration. Notably, despite differences in statistical significance, the residual associations between self-reported and device assessed sleep duration and internalizing problems were very similar (Supple. Table 4 and Supple. Table 5). In the other sensitivity analysis restricting sleep duration to a range of 7 to 10 hours (Supple. Figure 6), we found stronger phenotypic associations and similar genetic confounding effects. Sex-stratified results in this analysis found no clear evidence for genetic confounding (Supple. Table 6).

## Discussion

We conducted this study using both self-reported and device-based measures to compare differences in the associations of subjective and objective assessments of lifestyle behaviors with internalizing problems, as well as the genetic confounding, among adolescents. Genetic factor related to internalizing problems explained a substantial proportion of the association between self-reported sleep duration and internalizing problems. This genetic confounding was more evident among boys. Less clear evidence of genetic confounding was found in the association between device-measured sleep duration and internalizing problems. We found no evidence that the association between physical activity and internalizing symptoms was explained by shared genetic factors.

Our study identified genetic confounding in the association between self-reported sleep duration and internalizing problems. This identified share genetic risk may be partially explained by the shared method variance bias introduced by the questionnaire assessments for both exposures and outcome, as genetic confounding was observed more clearly with self-reported than device-based measures. This may be possibly due to the influence of mood-related genetic factors on sleep reporting.^44^ Adolescents of lower genetic risk may be more likely to overestimate their sleep duration when reporting and are less likely to develop internalizing problems compared to those of higher genetic risks. Yet, the sensitivity analyses using depression PGS revealed that genetic confounding in both sleep duration assessments were present among boys. Therefore, these results may also be explained by the shared genetic mechanisms that contribute to biological pathways that underlie both sleep and the broader internalizing problems. This can be evidenced by significant genetic correlations between insomnia and depression (genetic correlation of 0.41), and an extensive genetic overlap between sleep duration and depression.^45,46^ Previous research has highlighted the genetic link between sleep duration and depression, with several depression-associated genes also impacting sleep duration.^47^

In contrast, the lack of genetic confounding in physical activity suggests that mood-related genetic factors have less influence on reported physical activity levels than on sleep duration. This finding implies that device-based assessments may reduce the overestimation of associations between certain lifestyle factors and mental health only. Additionally, it is possible that there are only few shared genetic factors between physical activity and internalizing problems. This is consistent with a previous study showing that sleep duration has higher genetic correlation with depression than moderate/vigorous physical activity.^48^ Also, we might have had insufficient power from the GWAS of physical activity to accurately detect such smaller confounding effects.

Interestingly, the residual associations between sleep duration and internalizing problems were very similar across the assessment methods, after controlling for genetic confounding using depression PGS. This provides further evidence that bias by self-reporting (i.e., shared method variance) may be captured by the genetic confounding, although other biases might remain unaccounted. Even though no genetic confounding was found between either measure of physical activity and internalizing problems, self-reported physical activity showed a stronger association with internalizing problems than device-based measured physical activity. This difference may be explained by the shared method variance. However, this would likely be unrelated to any genetic predisposition to mood problems but influenced by other factors such as social desirability or cognition.^49^ Additionally, the accuracy of Fitbit in measuring moderate to vigorous physical activity may be less reliable compared to other accelerometer measures, introducing potential non-differential information bias.^50,51^ Finally, it may be contributed by the difference in measurements that self-reported activity reflects routine and intended physical activities, while the device-based measure includes actual activities, such as intensive outdoor play. This suggests that a lack of routine and intentional exercise is more closely related with increased internalizing problems than recreational or spontaneous activities.

We found a significant residual association between adolescent sleep duration and internalizing problems after accounting for genetic confounding among girls. This association was consistent across different assessment methods suggesting that longer sleep duration is beneficial for improving internalizing problems, especially for girls. This aligns with previous longitudinal and Mendelian randomization studies showing a potentially causal association between insomnia and depression.^52-55^ Additionally, higher physical activity levels were associated with less internalizing problems, even after accounting for genetic confounding.

Again, these findings are consistent with previous longitudinal and Mendelian randomization studies,^56-59^ although other behavioral or environmental factors, such as sedentary behavior while watching screens and parenting styles, may confound associations with physical activity.^60,61^ The results highlight the potential benefits of longer sleep duration and more moderate to vigorous physical activity for adolescents mental health. Although we should be cautious about other potential environmental confounders that were not considered in our study, such as cognition or sedentary activity level.

Our sex-stratified analyses of internalizing PGS relating with child lifestyle factors and internalizing problems revealed sex differences in the genetic confounding effects, although these differences were not statistically significant. The sensitivity analyses using depression PGS showed more pronounced sex differences. This is consistent with a whole-exome sequencing association study showing higher genetic burden of protein-truncating and deleterious variants for depression in males.^62^ Additionally, the genetic confounding explained part of the shared method variance, and this may be more prominent in boys. Previously, it has been reported that men tended to overreport their sleep time whereas women were more likely to report sleep duration accurately and much more in line with data obtained by a device.^63,64^

This study has several limitations. First, the association between sleep duration and internalizing problems may be non-linear, while our model assumes a linear relationship. However, sensitivity analyses suggest that the non-linearity has only a modest effect on genetic confounding. Second, while we adjusted for socio-demographic variables, other non-genetic confounding factors may still influence the associations between lifestyle factors and internalizing problems. Third, we focused on depression-related genetic factors and did not include genetic components related to sleep and physical activity due to limitations in the power of PGS derived from GWAS with smaller sample sizes.

## Conclusion

This study, to our knowledge, is the first to explore the role of shared genetic risk factors in the association between potentially modifiable lifestyle factors and internalizing problems. We show that the association of sleep duration with internalizing problems was more likely to be affected by genetic confounding, in particular if both exposure and outcome were assessed by self-report. The same genetic risk factors may underlie both sleep duration and internalizing problems. Additionally, genetic factors may explain reporting tendencies and constitute shared method variance bias that inflates associations. In contrast, the association between physical activity and internalizing problems was robust to genetic cofounding, whether physical activity was measured by self-reported questionnaire or objectively assessed by a device. These insights underscore the importance of considering shared genetic risks in socio-behavioral studies of lifestyle factors and youth mental health, especially studies relying on questionnaire-based measures alone that may be vulnerable to genetic confounding.

## Data Availability

All data produced in the present study are available upon reasonable request to the authors.

## Notes

### Competing Interest Statement

The authors have declared no competing interest.

### Funding Statement

Dr. Choi was supported in part by funding from the National Institute of Mental Health (K08MH127413) and a Hood Foundation Child Health Research Award. Dr. Pingault reported receiving grants from the European Research Council European Union Horizon 2020 Research and Innovation Programme during the conduct of the study. No other disclosures were reported.

### Author Declarations

The study used ONLY openly available human data that were originally located at National Institute of Mental Health Data Archive for Adolescents Brain Cognitive and Development cohort data release 5.0 (https://nda.nih.gov/study.html?id=2147).

