## Supplemental materials for "The association of objectively and subjectively measured *modifiable* lifestyle factors with internalizing problems: The role of genetic confounding and shared method variance *bias*"

### Supplemental Methods

The internalizing problems mean scores were calculated based on 6 items included in the Brief Problem Monitor instrument reported by children at their 13-year-old. The 6 items were 1) I feel worthless or inferior (Definition of inferior: less good); 2) I am too fearful or anxious; 3) I feel too guilty; 4) I am self-conscious or easily embarrassed; 5) I am unhappy, sad, or depressed; 6) I worry a lot. The mean will be calculated when at least 4 items were scored.

**Supplemental Table 1.** Demographic summary among analytical samples and overall European samples in the ABCD cohort.

| | Sleep analytical sample (N=2253) | PA analytical sample (N=2772) | Overall European (N=8795) |
|---|---|---|---|
| **Age at 2-year Follow-up (in years)** | | | |
| Mean (SD) | 12 (0.65) | 11.9 (0.65) | 12 (0.67) |
| Missing | 0 (0%) | 0 (0%) | 540 (6.1%) |
| **Sex at birth** | | | |
| Male | 1176 (52.2%) | 1457 (52.6%) | 4639 (52.6%) |
| Female | 1077 (47.8%) | 1315 (47.4%) | 4156 (47.1%) |
| **Parental Education level** | | | |
| High School Graduation | 431 (19.1%) | 553 (19.9%) | 2722 (30.9%) |
| Bachelor's degree | 734 (32.6%) | 886 (32.0%) | 2542 (28.8%) |
| Graduate Degree | 1087 (48.2%) | 1332 (48.1%) | 3526 (40.0%) |
| Missing | 1 (0.0%) | 1 (0.0%) | 5 (0.1%) |
| **Family income level** | | | |
| Less than $50,000 | 209 (9.3%) | 269 (9.7%) | 1669 (18.9%) |
| $50,000 - $100,000 | 652 (28.9%) | 866 (31.2%) | 2481 (28.1%) |
| More than $100,000 | 1298 (57.6%) | 1530 (55.2%) | 4103 (46.5%) |
| Missing | 94 (4.2%) | 107 (3.9%) | 542(6.2%) |
| **Sleep duration (hours) or physical activity frequency (days/week)** | | | |
| Self-reported | 9.3 (1.0) | 4.1 (2.1) | NA |
| Fitbit-measured | 7.6 (0.5) | 1.5 (1.7) | NA |
| **Mean Fitbit-measured time (days)** | 14.9 (9.3) | 12.5 (8.4) | NA |
| **Internalizing problems at 3-year Follow-up (mean scores)** | | | |
| Mean (SD) | 0.34 (0.39) | 0.34 (0.40) | 0.35 (0.41) |
| Missing | 67 (3.0%) | 89 (3.2%) | 961 (10.9%) |

**Supplemental Figure 1.** Sex-stratified phenotypic associations between modifiable risk factors and internalizing problems reported by children.

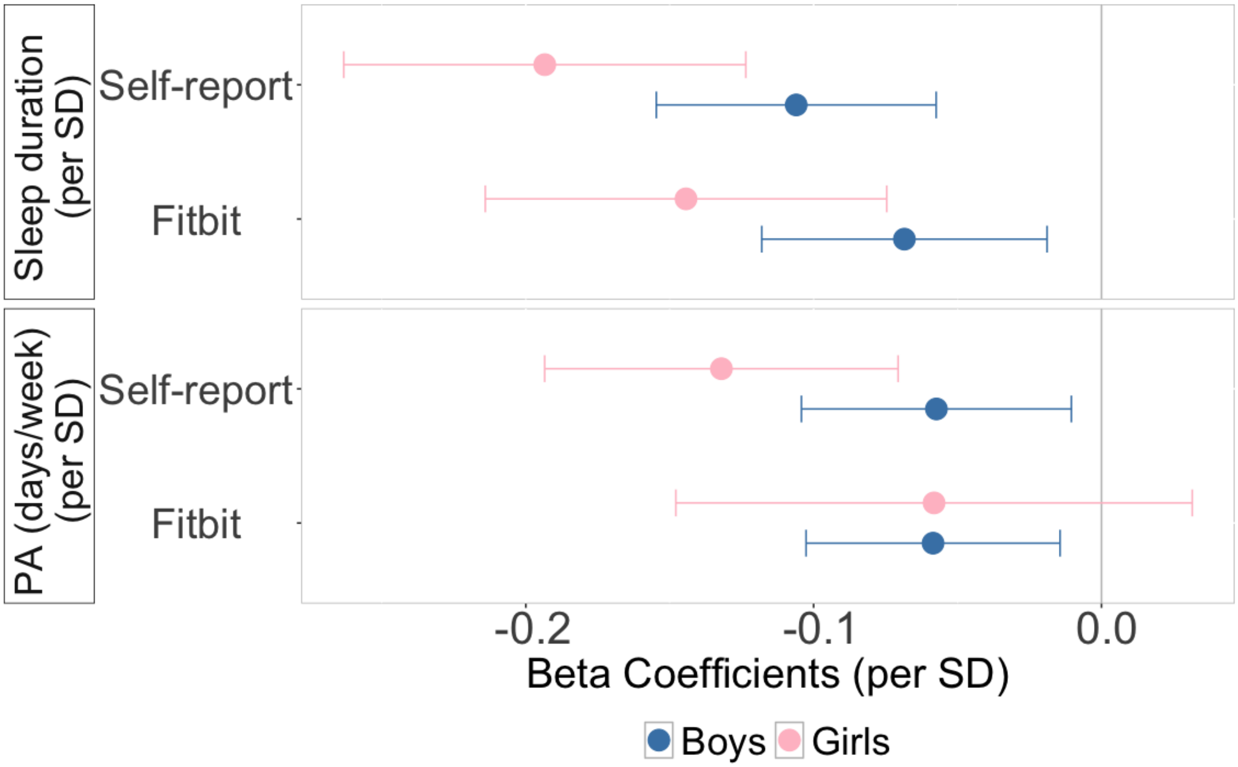


All models were adjusted for sex, age, family income and parental education, clustered with study sites.

**Supplemental Figure 2.** Phenotypic associations between modifiable risk factors and internalizing problems reported by parents.

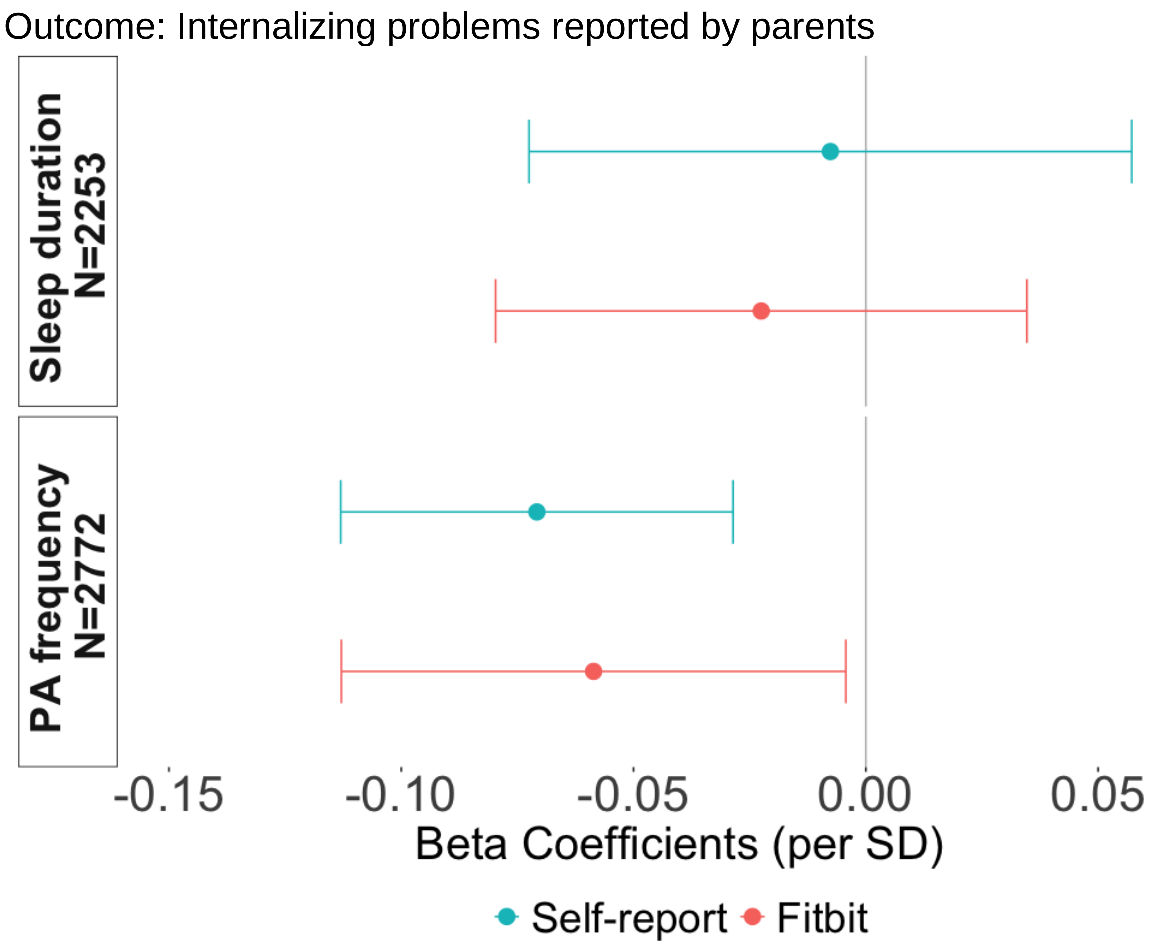

**Supplemental Figure 3.** Depression PGS associations with modifiable risk factors and internalizing problems.

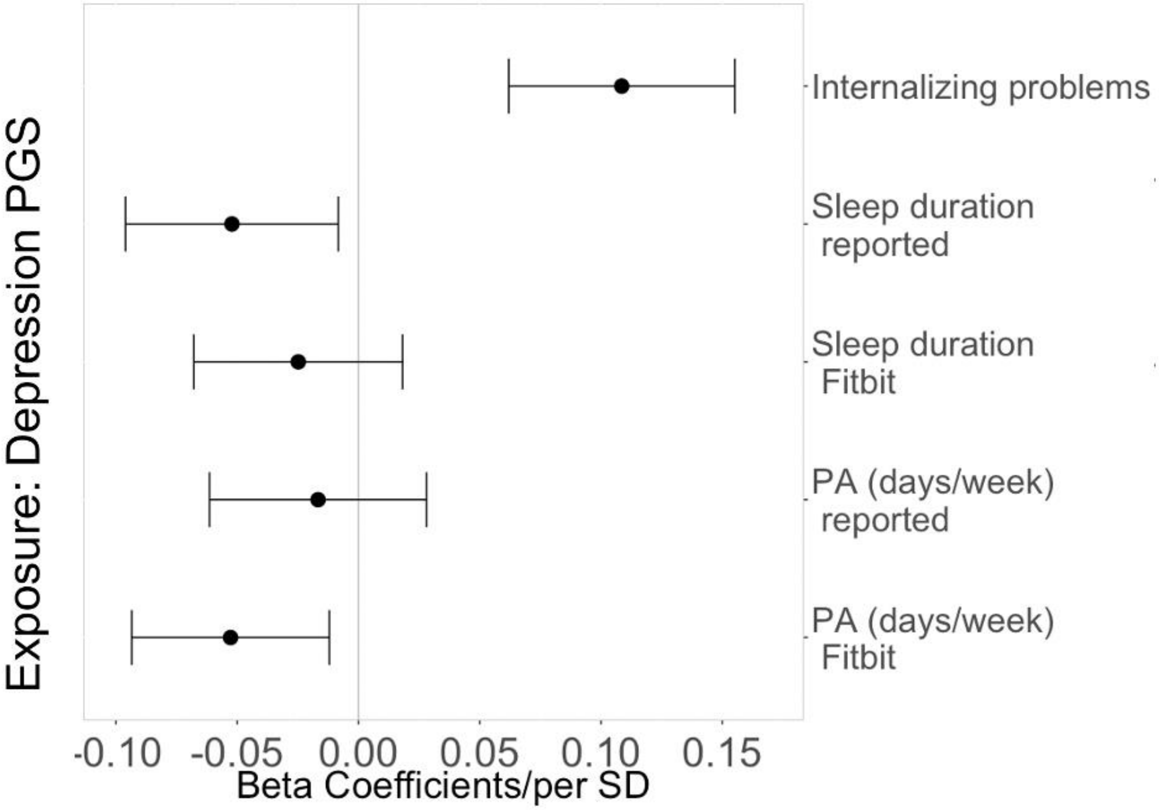


All models were adjusted for age, top 10 PCs and clustered with study sites. Note: The analytical sample is the overlapping sample between sleep and physical activity samples (N = 2084).

**Supplemental Figure 4.** Sex-stratified internalizing PGS associations with modifiable risk factors and internalizing problems

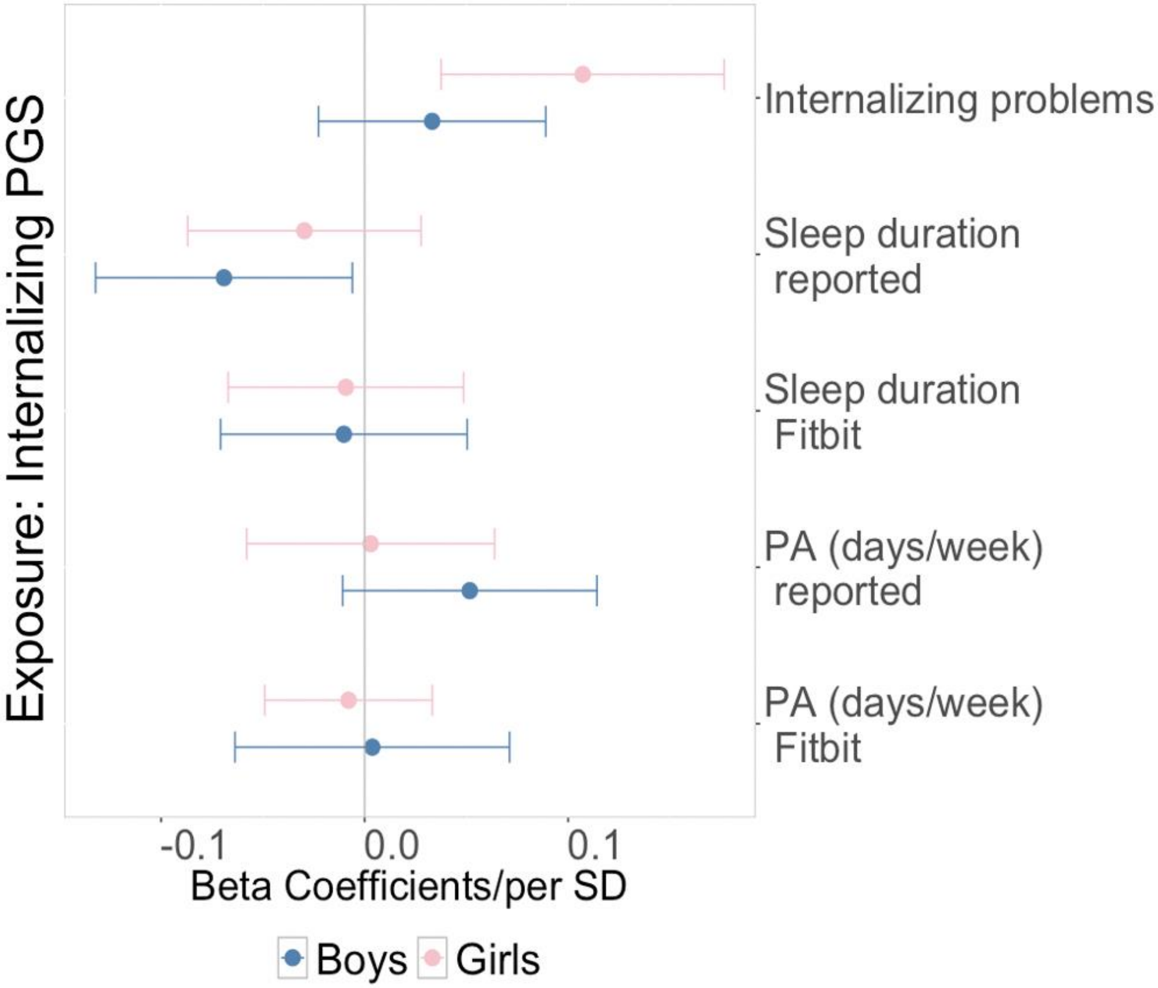


All models were adjusted for age, top 10 PCs and clustered with study sites. Note: The analytical sample is the overlapping sample between sleep and physical activity samples (N = 2084).

**Supplemental Figure 5.** Sex-stratified depression PGS associations with modifiable risk factors and internalizing problems

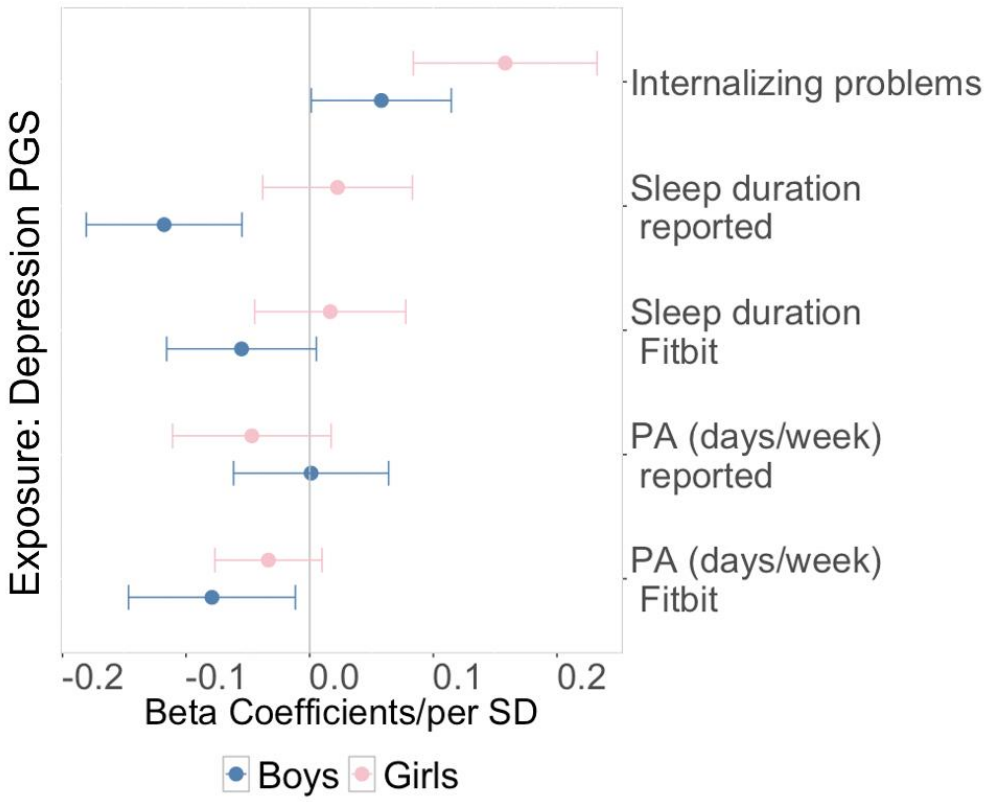

**Supplemental Table 2.** Demographic summary among sleep analytical samples and sex-stratified samples in the ABCD cohort.

| Sleep | **Overall sample (N=2253)** | **Girls (N=1077)** | **Boys (N=1176)** |
|---|---|---|---|
| **Age at 2-year Follow-up (in years)** | | | |
| Mean (SD) | 12 (0.65) | 12 (0.65) | 12 (0.65) |
| **Parental Education level** | | | |
| High School Graduation | 431 (19.1%) | 210 (19.5%) | 221 (18.8%) |
| Bachelor's degree | 734 (32.6%) | 349 (32.4%) | 385 (32.7%) |
| Graduate Degree | 1087 (48.2%) | 518 (48.1%) | 569 (48.4%) |
| Missing | 1 (0.0%) | 1 (0.1%) | 0 (0%) |
| **Family income level** | | | |
| Less than $50,000 | 209 (9.3%) | 103 (9.6%) | 106 (9.0%) |
| $50,000 - $100,000 | 652 (28.9%) | 326 (30.3%) | 326 (27.7%) |
| More than $100,000 | 1298 (57.6%) | 599 (55.6%) | 699 (59.4%) |
| Missing | 94 (4.2%) | 49 (4.5%) | 45 (3.8%) |
| **Sleep duration (hours)** | | | |
| Self-reported | 9.32 (1.0) | 9.33 (1.0) | 9.31 (0.9) |
| Fitbit-measured | 7.65 (0.5) | 7.71 (0.5) | 7.58 (0.5) |
| **Mean Fitbit-measured time (days)** | 14.9 (9.3) | 15.2 (10.3) | 14.6 (8.4) |
| **Internalizing problems at 3-year Follow-up (mean scores)** | | | |
| Mean (SD) | 0.34 (0.39) | 0.44 (0.44) | 0.25 (0.32) |
| Missing | 67 (3.0%) | 30 (2.8%) | 37 (3.1%) |

**Supplemental Figure 6.** Scatter plot with scaled sleep and scaled internalizing problems

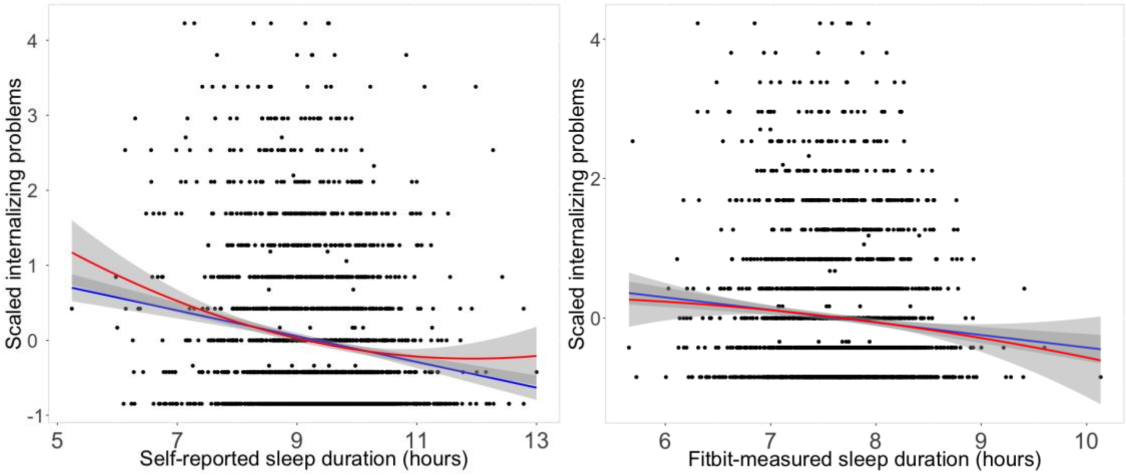


**Supplemental Table 3.** PGS of internalizing factor and depression associations with modifiable risk factors and internalizing problems.

| | Internalizing PGS | | Depression PGS | |
|---|---|---|---|---|
| | beta | 95% CI | beta | 95% CI |
| Internalizing problems at 3-year follow-up | 0.07 | 0.03, 0.12 | 0.11 | 0.06,0.16 |
| Self-reported sleep duration | -0.05 | -0.10, -0.01 | -0.05 | -0.10, -0.01 |
| Fitbit-measured sleep duration | -0.01 | -0.05, 0.03 | -0.03 | -0.07, 0.02 |
| Self-reported physical activity | 0.03 | -0.01, 0.07 | -0.02 | -0.06, 0.03 |
| Fitbit-measured physical activity | 0.001 | -0.386, 0.041 | -0.06 | -0.10, -0.02 |

**Supplemental Table 4.** Genetic confounding between child sleep duration and internalizing problems (sensitivity analyses using depression PGS).

| Reported Sleep – INT (N = 2253) | | | | Fitbit Sleep – INT (N = 2253) | | | |
|---|---|---|---|---|---|---|---|
| | beta | 95% CI | p | | beta | 95% CI | p |
| Total association | -0.147 | -0.188, -0.106 | <0.001 | Total association | -0.103 | -0.145, -0.062 | <0.001 |
| Residual association | -0.067 | -0.141, 0.008 | 0.08 | Residual association | -0.056 | -0.127, 0.015 | 0.12 |
| Genetic confounding | -0.080 | -0.144, -0.017 | 0.01 | Genetic confounding | -0.047 | -0.109, 0.014 | 0.13 |
| Reported Sleep – INT in **Boys** (N = 1176) | | | | Fitbit Sleep – INT in **Boys** (N = 1176) | | | |
| | beta | 95% CI | | | beta | 95% CI | p |
| Total association | -0.121 | -0.171, -0.71 | <0.001 | Total association | -0.073 | -0.126, -0.020 | 0.007 |
| Residual association | 0 | NA | NA | Residual association | 0 | NA | NA |
| Genetic confounding | -0.121 | -0.171, -0.71 | <0.001 | Genetic confounding | -0.073 | -0.126, -0.020 | 0.007 |
| Reported Sleep – INT in **Girls** (N = 1077) | | | | Fitbit Sleep – INT in **Girls** (N = 1077) | | | |
| | beta | 95% CI | p | | beta | 95% CI | p |
| Total association | -0.181 | -0.240, -0.122 | <0.001 | Total association | -0.138 | -0.197, -0.079 | <0.001 |
| Residual association | -0.165 | -0.280, -0.050 | 0.004 | Residual association | -0.117 | -0.232, -0.001 | 0.04 |
| Genetic confounding | -0.010 | -0.123, 0.091 | 0.77 | Genetic confounding | -0.021 | -0.128, 0.086 | 0.70 |

Note: depression PGS and internalizing problem heritability were used in the models. PGS was residualized on age, sex, top 10 PCs and clustered with study sites, and exposures were residualized on age, sex, family income, parental education and clustered with study sites.

**Supplemental Table 5.** Genetic confounding between child physical activity and internalizing problems (sensitivity analyses using depression PGS).

| Reported PA – INT (N = 2772) | | | | Fitbit PA – INT (N = 2772) | | | |
|---|---|---|---|---|---|---|---|
| | beta | 95% CI | p | | beta | 95% CI | P |
| Total association | -0.100 | -0.137, -0.063 | <0.001 | Total association | -0.049 | -0.087, -0.012 | 0.01 |
| Residual association | -0.139 | -0.239, -0.039 | 0.006 | Residual association | -0.034 | -0.123, 0.056 | 0.46 |
| Genetic confounding | 0.039 | -0.056, 0.135 | 0.42 | Genetic confounding | -0.016 | -0.100, 0.068 | 0.71 |
| Reported PA – INT in **Boys** (N = 1176) | | | | Fitbit PA – INT in **Boys** (N = 1176) | | | |
| | beta | 95% CI | | | beta | 95% CI | p |
| Total association | -0.074 | -0.132, -0.016 | 0.01 | Total association | -0.074 | -0.131, -0.017 | 0.01 |
| Residual association | -0.064 | -0.183, 0.055 | 0.29 | Residual association | 0.015 | -0.141, 0.171 | 0.85 |
| Genetic confounding | -0.010 | -0.118, 0.098 | 0.85 | Genetic confounding | -0.089 | -0.234, 0.056 | 0.23 |
| Reported PA – INT in **Girls** (N = 1077) | | | | Fitbit PA – INT in **Girls** (N = 1077) | | | |
| | beta | 95% CI | p | | beta | 95% CI | p |
| Total association | -0.125 | -0.184, -0.066 | <0.001 | Total association | -0.034 | -0.094, 0.025 | 0.26 |
| Residual association | -0.066 | -0.188, 0.056 | 0.29 | Residual association | \ | \ | \ |
| Genetic confounding | -0.059 | -0.172, 0.055 | 0.31 | Genetic confounding | \ | \ | \ |

PGS was residualized on age, sex, top 10 PCs and clustered with study sites, and exposures were residualized on age, sex, family income, parental education and clustered with study sites.

**Supplemental Table 6.** Sensitivity analysis for genetic confounding between self-reported sleep duration (between 7 and10 hours) and internalizing problems.

| Reported Sleep 7-10 hours – INT (N = 1712) | | | |
|---|---|---|---|
| | beta | 95% CI | p |
| Total association | -0.166 | -0.214, -0.119 | <0.001 |
| Residual association | -0.050 | -0.168, 0.068 | 0.41 |
| Genetic confounding | -0.116 | -0.223, -0.009 | 0.03 |
| Reported Sleep – INT in **Boys** (N = 893) | | | |
| | beta | 95% CI | |
| Total association | -0.151 | -0.216, -0.085 | <0.001 |
| Residual association | -0.040 | -0.238, 0.157 | 0.69 |
| Genetic confounding | -0.110 | -0.294, 0.073 | 0.24 |
| Reported Sleep – INT in **Girls** (N = 819) | | | |
| | beta | 95% CI | |
| Total association | -0.182 | -0.251, -0.112 | <0.001 |
| Residual association | -0.015 | -0.228, 0.198 | 0.89 |
| Genetic confounding | -0.167 | -0.368, 0.035 | 0.10 |

PGS was residualized on age, sex, top 10 PCs and clustered with study sites, and exposures were residualized on age, sex, family income, parental education and clustered with study sites.
